# Test-Retest Reliability of Assessment of Work Performance for Thai Homeless People

**DOI:** 10.1101/2025.01.23.25321059

**Authors:** Winai Chatthong, Uthaikan Thanapet, Watthanaree Ammawat, Maliwan Rueankam, Supalak Khemthong

**Affiliations:** Mahidol University

**Keywords:** homeless, mental illness, vocational rehabilitation, work performance assessment, test-retest reliability

## Abstract

**Purpose:** This study aims to conduct test-retest reliability of the AWP – Thai version. Internal consistency has been tested after completing an approval process of forward-backward translation and achieving good content validity. Under the shortage of mental workforce, vocational rehabilitation (VR) has been trained to enhance supported employment in homeless people with mental illness (MI). The Assessment of Work Performance (AWP) is required for Thai translation to classify the quality of job skills in one pilot setting at Nonthaburi’s destitute home.

**Methods:** Two weeks were set for test-retest reliability of the AWP collecting a group of participants (n = 59). Intraclass Correlation Coefficient (ICC) was calculated at the 95% confidential level with an F-test of true value.

**Results:** Moderate to good test-retest reliability of the AWP is addressed for all participants and separation of those participants into four groups of job selection: rug hooking, cleaning food table, sweeping the leaves, and community gardening. However, a cautious interpretation of ICC provides accurate implications of the AWP in both clinical utility and the VR program evaluation.

**Conclusions:** The AWP is a performance-based tool that can be an observable process in participatory work routines. Motor, process, and interactive communication skills are essential components connected to a practical solution to enhance the VR to the social welfare system.

## Introduction

Vocational Rehabilitation (VR) has been trained for 3 years by psychosocial occupational therapists among 114 caregivers working at 11 Thai destitute homes. Overloaded caregiving services were found at a ratio of one caregiver to 148-371 homeless people with mental illness (MI). A needs assessment was then conducted among those MI clients (n = 3,201); their functional abilities were partially performed on self-care activities (27.31%), maintained intrinsic capacity for productivity (10.08%), and needed further assessment of work performance (62.60%).

Previous pieces of evidence highlight that an integrated knowledge exchange strategy (IKE) is an important concept for interprofessional collaborative development [1] in parallel to a peer-support worker advisory group including a social justice engagement [2]. Pre- and post-evaluation is also a way of understanding how interdisciplinary collaboration works through a cross-training model [2]. This model seems to be helpful lessons learned from situational analysis and synthesis of practical solutions in any field of mental health rehabilitation areas such as positional clarification, modeling, and role rotation in the same case.

A progressive sense of the VR training was determined into three rehabilitation principles [3] among those caregivers: purposeful relationship to meaningful nourishing, healing cultures with careful leadership, and compassionate communication-based practical solutions leading to Mental Health Recovery (MHR) workers like supported employment [4]. Importantly, a semi-structured observation during work performance may increase MHR among people with both MI and restrictive issues with the social welfare system [5].

Consequently, community reintegration of people with mental health problems seems to be an innovative collaboration among MHR workers working in a transitional program between a prolonged stay of IPD and community rehabilitation [6]. Unfortunately, if there is no prolonged stay IPD for the government mental health hospitals in Thailand then all Thai destitute homes must be long-stay rehabilitation despite lacking occupational therapists. For a standard of practice [6], a weekly or 2-weekly teleconference must be conducted during the transitional process before the hospitalized discharge from a long-stay rehabilitation between 6-8 weeks (7 nights per week). A sense of progression is emphasized to see ‘What works are in any supported learning community with no right or wrong’[7]. However, homeless people with MI need to promote purposeful decision-making on the VR with homeless people with MI, an assessment of work performance or AWP is practically required.

The AWP was developed by Sandqvist et al. in 2006 as an observation-based assessment tool that allows using individuals’ actual work tasks [8]. This comprehensive assessment enables ordinal scorings of three domains: physical, process, and communication/integration skills. To accomplish supported employment the VR of individualized task preferences is suggested during the AWP [9]. Also, an individualized reskilling plan can be designed after the AWP is scored together with identifying personal strengths and limitations [10]. The AWP is conceptualized using the Model of Human Occupation (MOHO) which provides occupation-focused outcomes in structural working activities and realistic life engagement [11, 12]. However, this is the first reliable testing of the AWP among homeless people with MI at one destitute home in Thailand. Hopefully, this study will enable the Thai version of AWP for occupational therapists and MHR workers related to VR settings in the future.

## Methods

### Design

This cross-sectional test-retest reliability study was conducted. Interprofessional collaboration was communicated among social workers, psychologists, occupational therapy students, and academics. A permission agreement contract was made to translate the AWP to Thai (version 2.0) between Unitalent representing Jan Sandqvist and the corresponding author on 15 Dec 2022.

### Participants

Purposive samplings were used among 212 homeless people who were classified independent level of self-care abilities living at Nonthaburi’s destitute home. The interview protocol contains three parts. Part A included questions on the demographics of the participants. Part B comprised a cut-off equal to or more than 23 scores of the Rowland Universal Dementia Assessment (RUDAS) and a cut-off equal to or less than 36 scores of the Brief Psychiatric Rating Scale (BPRS). Part C consisted of a checklist of inclusion criteria: (1) be over 18 years old and no later than 64 years, (2) born in Thailand and used the Thai language; (3) be diagnosed with MI – no delusion and acute episode, (4) have no physical handicap, (5) Able to use both hands – no neurological deficits, and (6) have no low vision/deafness. All sample sizes (n = 60) have been calculated using the statistical power at 80% with a confidential level of 95% to protect against bias and to achieve acceptable overall reliability [13].

### Measurement

A sociodemographic form was used in this study including age, gender, education, diagnosis, experienced drug addiction, received non-chronic disease/s, and status of the VR. The AWP (Thai version) was also implemented after its forward-backward translation and content validity was acceptable agreement. Although a specific task was not required for the AWP, those homeless participants continued undergoing the VR: two-month supported employment and in-house pre-vocational training. The AWP was designed to assess three skills of work performance: 5 items of motor skills, 5 items of process skills, and 4 items of communication/interaction skills. Quality of work performance was observed and noted in terms of efficiency, adequacy, and appropriateness on its blank box. Scoring was rated per one item using a four-point ordinal scale (1 = deficient performance, 2 = inefficient performance, 3 = uncertain performance, 4 = competent performance). If an assessor did not know adequate information or relevant situations to rate each item in the client’s working context, the alternative notes would be lack of information (LI) and not relevant (NR) respectively. The total administration time of rating the 14-item AWP was a flexible observation, depending upon the working situation of individuals. When the participants were noted as LI or NR, the summation of the total score could not be completely computed [12].

### Translation and content validity

The corresponding author translated an original English version of AWP into Thai. Then, a bilingual person translated the Thai version into English (the 2^nd^ draft). After that Jan Sandqvist approved the Thai version 2.0 of AWP on 29 August 2023. The content validity index, or CVI would be accepted if accounting no less than 0.8 for all items from three experts in psychosocial occupational therapy. Some useful comments from those experts were brought to correct all keywords becoming the Thai version 3.0 of AWP which was applied to those participants.

### Job selection

Before the first round of data collection, the researchers asked one psychologist and one social worker for job selection to be observed in the AWP. The existing two-month VR programs were selected: supported employment as community gardening and in-house pre-vocational training i.e., rug hooking in a tutorial group, cleaning tables in a food corner, and sweeping the leaves/growing small plants. Six occupational therapy students were assigned to observe and score those participants while doing the VR programs for 6 consecutive days.

### Data Analyses

Statistically, IBM SPSS V.22 was used and Cronbach’s alpha for the three domain scores and the total score of AWP were calculated considering an acceptable level of reliability from 0.60 to 0.70 and an excellent level from 0.8 to 0.95 [14]. The corrected items of the AWP were rated during personal observation twice in two-week intervals based on a wash-out time. Intraclass Correlation Coefficient (ICC) was calculated using two-way mixed effects of test-retest reliability, and absolute agreement of single rater. A 95% confidence interval has been suggested, with values of 0.50 or higher indicating moderate agreement and values of 0.75 or higher indicating good agreement [15].

## Results

Among 212 homeless people, the participants (n = 72) were willing to engage both screening tools of the RUDAS and the BPRS. Sixty participants met the inclusion criteria, but only one male was withdrawn because of no attendance in the second assessment (Table 1). Mean (SD) of those participants (n = 59) aged 44.19 (7.72) and lived in the Nonthaburi’s destitute home 52.71 (37.83) months. Most participants graduated from primary school and experienced unspecified mental disorders as well as drug addiction. Community gardening was the highest percentage for vocational rehabilitation among those participants.

**Table 1.**
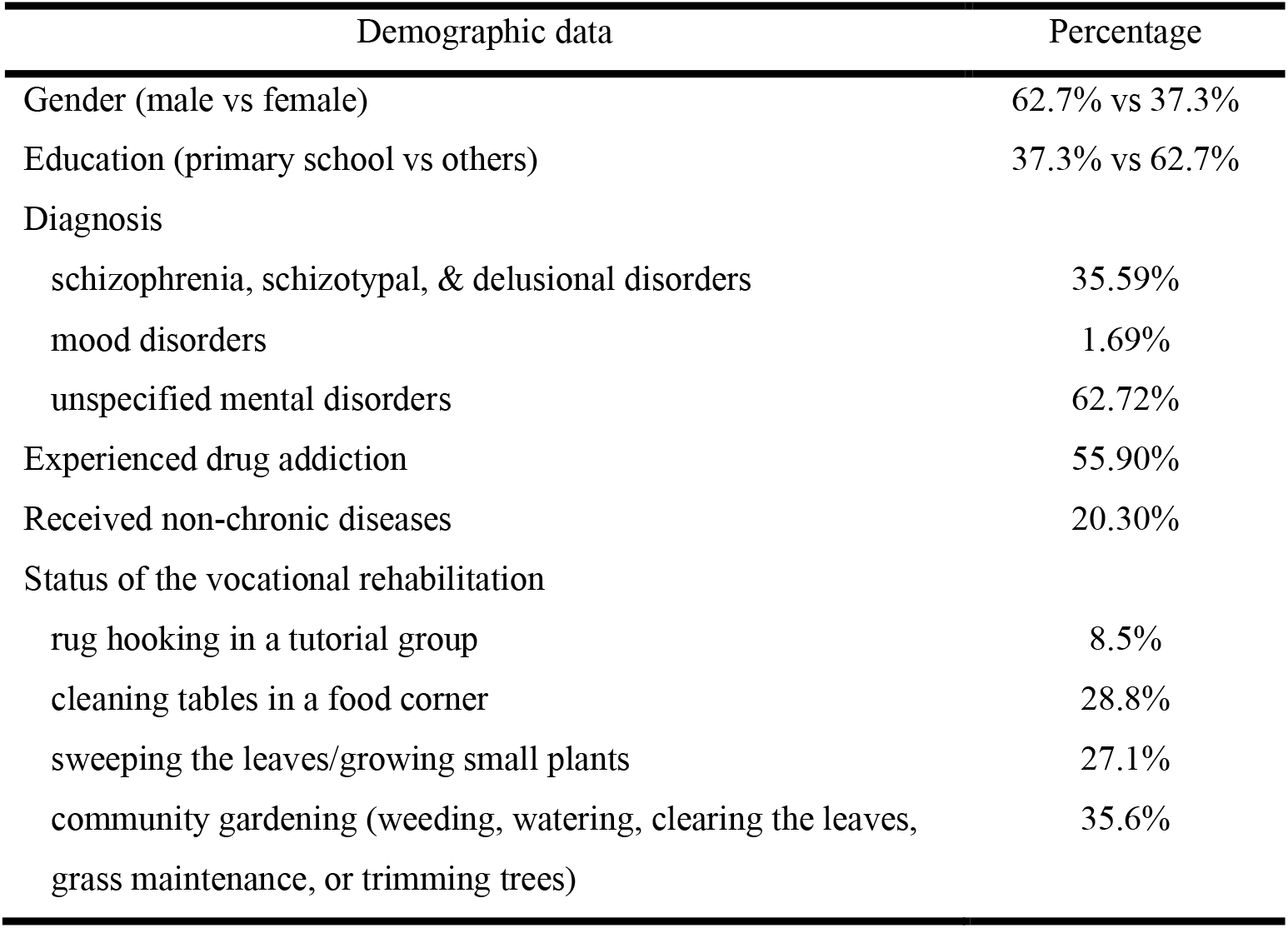
Demographic percentage of the participants (n = 59)

Three experts’ content validity was determined for acceptable language in the Thai context with a CVI of 1.0 0. Fourteen items were finally scored for the first AWP (n = 59) to achieve Cronbach’s alpha at 0.74 (good internal reliability). As seen in Table 2, the participants performed their roles and responsibilities by taking a few steps of similar tasks and assessors. The ICC of test-retest reliability was significantly found; to moderate reliability with an F-test of true value at the 95% confidential level from 0.60 to 0.77.

**Table 2.** Descriptive mean (SD) of AWP scores.

| AWP scores | The first<br>assessment | The second assessment | ICC<br>n = 59 |
| --- | --- | --- | --- |
| Motor skills | 19.58 (0.83) | 19.68 (0.80) | 0.64 |
| Process skills | 18.59 (1.91) | 19.17 (1.09) | 0.60 |
| Communication<br>& interaction skills | 14.47 (1.64) | 14.80 (1.35) | 0.78 |
| Total score | 52.59 (3.25) | 53.64 (2.48) | 0.77 |

*SD* standard deviation; *AWP* assessment of work performance; *ICC* interclass correlation coefficient.

When the ICC was made to determine three domains and summation of the AWP scores there were four separate groups to be compared (Table 3). All domains and the total test demonstrated moderate to good test-retest reliability (ICC = 0.59-0.85) while the participants performed cleaning tables in a food corner (n = 17). The AWP of communication and interaction skills showed moderate to excellent test-retest reliability (ICC = 0.63-0.95) across the four job selections. However, there were non-significant F-tests of true value (p > .05): motor skills in rug hooking and gardening in-house training, and process skills in rug hooking and sweeping leaves/growing small plants.

**Table 3.** Comparison of test-retest reliability for different tasks of AWP.

| AWP scores | ICC<br>rug hooking<br>n = 5 | ICC<br>cleaning<br>n = 17 | ICC<br>sweeping<br>n = 16 | ICC<br>gardening<br>n = 21 |
| --- | --- | --- | --- | --- |
| Motor skills | NS | 0.72* | 0.78* | NS |
| Process skills | NS | 0.59* | NS | 0.66* |
| Communication<br>& interaction skills | 0.95* | 0.85** | 0.66* | 0.63* |
| Total score | 0.96* | 0.72* | 0.78* | 0.63* |

\* $p < .05$ ; \*\* $p < .001$ ; AWP, assessment of work performance; ICC, interclass correlation coefficient; NS, non-significant of true value.

## Discussion

This pilot study reports that three domains and total scores of the AWP enable observable work performance among Thai homeless people with MI. Sound content validity and acceptable internal consistency reliability are addressed. Although this participatory assessment seems to evaluate the working skills of individuals, job selection in either single-tasking or multi-tasking is a cautious interpretation. Occupational therapy students and academics can explain to caregivers and multi-disciplinary how those participants have been intrinsically motivated and progressively performed motor, process, and interactive communication skills. This statement is supported by the previous pieces of evidence [2, 3, 7]. Since the AWP has been created according to the MOHO [8] occupational therapists and trained assessors may bring the AWP to the clinical utility of the Thai context for the VR outcome measurement, especially for homeless people with MI in chronic conditions. The current study is supported by Davutoglu and colleagues in 2023 [16], the AWP’s job selection is modified for an appropriate determination when the assessors have understood operational definitions of the 14-item components and the four-point ordinal scale including LI and NR. Each participant took a role with different tasks and flexible time, no specific task is required for volitional participation with individual preferences – the selected task is not too difficult or easy to perform [11, 12].

However, our Cronbach’s alpha of the AWP (Thai version) shows lower than the internal consistency of the original AWP of Sandqvist and colleagues [12] and the Turkish version [16]. This might be respondents’ external factors influencing the powerful significance of Cronbach’s alpha [17]; a higher noise and a lower education sense the lower value of Cronbach’s alpha. Different degrees of volition and habituation are found to be reconsidered including levels of interest, a wide range of age, and unequal numbers between feminine and masculine groups. Importantly, a previous study emphasized the scale anxiety-technological problem modifies the under 0.70 value of Cronbach’s alpha[17]. While a face-to-face situation is being undertaken some participants have perceived a circumstance of no existing reward, for instance, a paper-and-pencil interviewing, a simulated observation. The current study did notice one item of motor skills to be deleted to gain a 0.70 value of Cronbach’s alpha instead of 0.68 (14 items), however, the authors decided to keep all items of the AWP–Thai version for those vulnerable participants (n = 59) based on the good internal reliability [14] which was in contrast to a sample size of 60 participants suggested by Bujang & Baharum [13].

To imply the good test-retest reliability of the AWP (n = 59), this current study addresses an important approach for twice assessment to confirm no change in personalized work performance over two weeks. Meaningfully, a group of homeless people with MI would have received positive feedback through the utilization of the AWP before personalized planning (e.g., strengths, needs for upskilling/reskilling, how to maintain or maximize abilities) for supported employment or pre-vocational training at the destitute home. The authors are also interested in looking for different test-retest reliability of the AWP. All participants were grouped by asking about their routine of work assignments in the destitute home, three jobs were planned as simulated vocational training (i.e. in-house sweeping/planting, cleaning food table, and rug hooking) and only one job was daily asked for paid employment by an outside agency of metropolitan gardening.

Additionally, measuring an observational work performance is challenging unless a standardized task is created by understanding a task selection process to score the structured manner of individuals in a group-based task [16]. For example, a comprehensive skill set includes hand functions, coordination, executive functioning, and seeking help. the current study set a group-based tutorial activity for rug hooking toward self-controlled productivity. All participants (n = 5) attempted to complete one piece of rug for a person, a ceiling effect of ICC > 0.90 reflected a full score of more than 15% of total clients according to a useful suggestion of Fan and colleagues (2013). A high item separation score (> 3) can also classify one competent working skill domain of at least 10 clients for each job section process. A few tasks consisted of cleaning the food table (sweeping the floor, wiping the table, and collecting the dishes) related to three skill components of work performance: moderate reliability of process skills and good reliability of the other two domains including total score. Whereas multi-tasking in different periods of assessment time at community gardening the six assessors might observe a few out of 21 participants performed more than one task, for instance, grass maintenance in parallel to watering showed no significant ICC accounted for motor skills. Davutoglu and colleagues [16] suggest intensive job site training in a structured manner and constant observation between two assessors and one client. Needs for personal adaptation or supportive action should be considered in separated job selection from a sample size of 16-21 enabling moderate to good test-retest reliability.

## Limitations

Three issues are considered as our limitations. Firstly, most assessors are students, and this study lacks inter-rater reliability testing. To increase the sensitivity of the AWP, all assessors should follow the AWP manual guidelines and practice for achieving good inter-rater reliability. Secondly, this study uses the de-identified data without a cross-tabulation. Chi-square analysis will provide additional information related to personal variables that may be hidden from the AWP administration. Lastly, the variety of job selection processes with heterogeneous multitasking is a limitation of this current study. A focus-group assessment in pre- and post-training in the VR is further recommended analyzing purposeful and meaningful activity to be observed in natural situations. Internal consistency reliability is required to examine whenever a sample size per activity category and domain of the AWP is changed no later than 60 persons.

## Conclusions

Individuals with homeless and MI could be initially assessed using the Thai version of AWP. Before and after evaluation of work performance three domains of working skills are observable items. To enhance personal support for a further VR, the most important component is the job selection process in both prevocational training and supported employment. Inclusive training on how to shift a single task toward multitasking is concerned with high-quality assessors. If the AWP is combined with other vocational measurement tools, occupational therapists and relevant stakeholders will maintain the effectiveness of the VR and MHR process in the future. A caution of interpretative assessment outcome is suggested in different modes of MHR settings such as rehabilitation centers, community areas, and job coaching onsite.

## Data Availability

All data produced in the present study are available upon reasonable request to the authors

## Acknowledgments

The authors would thank occupational therapy students, academics, and experts. They would also like to thank Unitalent representing Jan Sandqvist for their permission agreement for the AWP translation and research. This project would not have been completed without support from all caregivers and clients at Nonthaburi’s destitute home.

## Author Contributions

All authors contributed to the study conception and design of the study. Material preparation and data collection was performed by Winai Chatthong, Uthaikan Thanapet, Watthanaree Ammawat and Maliwan Rueankam. Watthanaree Ammawat and Uthaikan Thanapet had formal analysis. Project administration and data curation were Maliwan Rueankam. The first draft of the manuscript was written by Supalak Khemthong and all authors commented on previous versions of the manuscript. Supalak Khemthong and Winai Chatthong had the main responsibility for writing and revising the manuscript. All authors read and approved the final manuscript.

## Funding

This project was made possible by funding for a new researcher at the Faculty of Physical Therapy, Mahidol University in the fiscal year 2024. Open access funding provided by Mahidol University.

## Declarations

### Conflict of interests

The authors declare that they have no conflict of interest.

### Ethics Approval

This study has received ethical approval from the Mahidol University Central Institutional Review Board (COE No. MU-CIRB 2023/186.2212). Permission to use consent information and assessment tools was considered for safety and confidentiality by the Department of Social Development and Welfare, Ministry of Social Development and Human Security, THAILAND.

### Informed Consent

Participants were fully informed and provided written consent to participate before conducting the assessment.

### Consent for Publication

All participants provided written consent to take part in the study and agreed that the results could be utilized for research purposes.

### Open Access

This article is available under the Creative Commons Attribution 4.0 International License, which allows for use, sharing, adaptation, distribution, and reproduction in any medium or format, provided that proper credit is given to the original author(s) and source, a link to the Creative Commons license is included, and any modifications are noted. Images or third-party content within this article are covered by the Creative Commons license, unless otherwise credited. If specific content is not covered by this license and your use goes beyond what is legally permitted, you must obtain permission from the copyright holder. The license can be viewed at http://creativecommons.org/licenses/by/4.0/.

## Notes

### Competing Interest Statement

The authors have declared no competing interest.

### Author Declarations

This study has received ethical approval from the Mahidol University Central Institutional Review Board (COE No. MU-CIRB 2023/186.2212)

### Summary of Updates

The explanation objective is written and the reconsideration about those confounding factors are described. The rationale and knowledge gap are edited. The contribution of this article is reconsidered and have been added to explain.The specific recruitment method is correctly written. The detail descriptions are written support these arguments.The details regarding the effectiveness of different measurement are added.The limitation of utilizing Test are newly written to take into consideration this practical work.

